# Assessment of functional capacity with cardiopulmonary exercise testing in non-severe COVID-19 patients at three months follow-up

**DOI:** 10.1101/2020.11.15.20231985

**Authors:** Piero Clavario, Vincenzo De Marzo, Roberta Lotti, Cristina Barbara, Annalisa Porcile, Carmelo Russo, Federica Beccaria, Marco Bonavia, Luigi Carlo Bottaro, Marta Caltabellotta, Flavia Chioni, Monica Santangelo, Arto J. Hautala, Pietro Ameri, Marco Canepa, Italo Porto

**Author notes:** **Address for correspondence** Prof. Italo Porto, University of Genoa, Cardiovascular Unit, Department of Internal Medicine and Specialties (DIMI) Viale Benedetto XV, 10, 16132 Genoa (Italy). These authors contributed equally to this work (Piero Clavario, Vincenzo De Marzo). **Declaration of interests:** none of the authors have conflict of interests for this work. **Funding sources:** none of the authors received funding for this work.

## Abstract

**Introduction:** Long-term effects of Coronavirus Disease of 2019 (COVID-19) and their sustainability in a large number of patients are of the utmost relevance. We aimed to determine: 1)functional capacity of non-severe COVID-19 survivors by cardiopulmonary exercise testing (CPET); 2)those characteristics associated with worse CPET performance.

**Methods:** We prospectively enrolled the first 150 consecutive subjects with laboratory-confirmed COVID-19 infection discharged alive from March to April 2020 at Azienda Sanitaria Locale (ASL)3, Genoa, Italy. At 3-month from hospital discharge, complete clinical evaluation, trans-thoracic echocardiography, cardiopulmonary exercise testing (CPET), pulmonary function test (PFT), and dominant leg extension (DLE) maximal strength evaluation were performed.

**Results:** Excluding severe and incomplete/missing cases, 110 patients were analyzed. Median percent predicted peak oxygen uptake (%pVO2) was 90.9(79.2-109.0)%. Thirty-eight(34.5%) patients had %pVO2 below, whereas 72(65.5%) above the 85% predicted value (indicating normality). Median PFT parameters were within normal limits.

Eight(21.1%) patients had a mainly respiratory, 9(23.7%) a mainly cardiac, 3(7.9%) a mixed-cardiopulmonary, and 18(47.4%) a non-cardiopulmonary limitation of exercise. Eighty-one(73.6%) patients experimented at least one symptom, without relationship with %pVO2 (p>0.05).

Multivariate linear regression analysis showed age (β=0.46, p=0.020), percent weight loss (β=-0.77, p=0.029), active smoke status (β=-7.07, p=0.019), length of hospital stay (β=-0.20, p=0.042), and DLE maximal strength (β=1.65, p=0.039) independently associated with %pVO2.

**Conclusions:** Half of non-severe COVID-19 survivors show functional capacity limitation mainly explained by muscular impairment, albeit cardiopulmonary causes are possible. These findings call for future research to identify patients at higher risk of long-term effects, that may benefit from careful surveillance and targeted rehabilitation.

**Take-home messages:** at 3-month cardiopulmonary exercise testing 38/110(34.5%) non-severe COVID-19 survivors had percent predicted peak oxygen uptake (%pVO2) < 85% (indicating normality). Half of them had functional capacity limitation mainly explained by muscular impairment.

## INTRODUCTION

To date, the Coronavirus Disease of 2019 (COVID-19) pandemic accounts for more than 50 million confirmed cases and up to 1 million deaths worldwide[1]. Whereas handling the initial phase has been challenging for health systems, the sheer number of cases raises alarm about the sustainability of even minor sequelae after hospital discharge.

COVID-19 is a mainly respiratory disease, but cardiovascular (CV) alterations are also associated with worse prognosis[2]. For the chronic phase, the main concerns are the development of pulmonary interstitial disease and/or a lingering CV involvement, as hypothesized by a recent CV magnetic resonance study[3] and potentially explained by the Severe Acute Respiratory Syndrome-Coronavirus type 2 (SARS-CoV2)-associated endothelitis[4]. How to intercept, assess, and treat this large number of patients with potential long-term consequences of COVID-19 remains uncertain, with the hypothesized rehabilitative effort mainly focused on the post-intensive care patients[5, 6]. It should be noted, however, that among the 20,000+ COVID-19 patients hospitalized in Italy and discharged alive, those classified as clinically severe or critical represent less than 20%[7].

Conversely, despite data from the 2003 SARS outbreak highlighting long-term exercise capacity reduction even in absence of cardiac or pulmonary abnormalities[8], data on long-term functional COVID-19 effects in less clinically complex patients are lacking.

Aims of our study were: 1) to evaluate pulmonary, cardiac, and functional capacity of non-severe COVID-19 survivors by performing cardio-pulmonary exercise testing (CPET); 2) to identify those baseline and clinical characteristics associated to worse performance at CPET.

## MATERIAL AND METHODS

### Study subjects

We included the first 150 consecutive subjects undergoing post-COVID-19 evaluation at the Outpatient Cardiac Rehabilitation center of Genoa, Italy. The local healthcare authority (Azienda Sanitaria Locale, ASL 3 Genovese) set up a structured follow-up program for all patients with a history of Reverse transcriptase-polymerase chain reaction (RT-PCR)-confirmed Severe Acute Respiratory Syndrome Coronavirus 2 (SARS-CoV-2) infection admitted to COVID-19 wards from 1^st^ of March 2020 to date (recruitment is still ongoing).

For the purpose of the study, we included in analysis all non-severe patients, excluding those requiring mechanical ventilation and intensive care.

### Study design

At 3 months from hospital discharge, all patients received complete clinical evaluation, trans-thoracic echocardiography (TTE), CPET, pulmonary function test (PFT), and dominant leg extension (DLE) maximal strength evaluation. All patients signed an informed consent.

Study protocol and informed consent conform to the Declaration of Helsinki and were approved by the Ethics Committee of the Liguria Region (n° 430/2020CER).

All procedures and protocols are described in detail in **online-only material**.

### Analysis

Categorical variables are presented as frequencies and percentages and were compared by chi-square test or Fisher’s exact test. Continuous variables are reported as mean and standard deviation (SD) or median and interquartile range (IQR) according to their distribution. Normally distributed variables were compared by means of unpaired Student’s t test. Non-normally distributed variables were compared with the U Mann-Whitney non-parametric test.

The main outcome measure was percent predicted peak VO2 (%pVO2). Patients were categorized according to the value of %pVO2 below or above 85%.

Multivariate linear regression model was used to estimate the beta coefficients with 95% confidence interval (CI) of %pVO2. The model was adjusted for time from hospital discharge to CPET and all clinically meaningful covariates with p <0.10 in univariate analysis.

All analyses were performed with R environment 3.6.3 (R Foundation for Statistical Computing, Vienna, Austria) and packages tableone, finalfit, and ggplot2.

## RESULTS

Of the first 150 evaluated patients, we excluded 24 (16.0%) who had needed invasive ventilation, 7 (4.7%) for missing data, and 9 (6.0%) as they were unable to perform CPET. The final population included 110 patients. **Table 1** depicts the characteristics of the study patients.

**Table 1.** **Characteristics of the study patients stratified according to percent predicted VO2 below/above 85%.**

| Variable | Overall population<br>(n=110) | % predicted VO2 below 85%<br>(n=38) | % predicted VO2 above 85%<br>(n=72) | p value |
| --- | --- | --- | --- | --- |
| **Baseline and clinical characteristics** |  |  |  |  |
| Age (years) | 61.7 (53.5-69.2) | 58.8 (51.4-67.7) | 62.4 (54.6-70.0) | 0.266 |
| Sex (female) | 45 (40.9) | 14 (36.8) | 31 (43.1) | 0.670 |
| Height (centimeters) | 168.0 (163.0-174.8) | 167.0 (163.0-173.0) | 169.0 (163.0-176.0) | 0.491 |
| Absolute weight loss (kg) | 7.5 (5.0-10.0) | 7.5 (6.0-12.0) | 7.5 (4.0-10.0) | 0.316 |
| Weight at CPET evaluation (kg) | 77.0 (67.0-89.8) | 72.5 (64.3-79.5) | 78.5 (69.5-90.3) | 0.010 |
| Weight at hospital admission (kg) | 82.0 (70.0-95.0) | 80.0 (66.0-85.5) | 83.0 (72.8-95.8) | 0.049 |
| Percent weight loss (%) | 9.4 (6.0-12.9) | 10.4 (7.6-15.6) | 9.2 (5.3-12.2) | 0.090 |
| Body mass index (kg/m<sup>2</sup>) | 26.8 (23.9-30.5) | 26.1 (23.1-29.2) | 28.0 (25.1-31.9) | 0.042 |
| Hypertension | 49 (44.5) | 19 (50.0) | 30 (41.7) | 0.525 |
| Diabetes | 3 (2.7) | 2 (5.3) | 1 (1.4) | 0.568 |
| Active smoke | 45 (40.9) | 18 (47.4) | 27 (37.5) | 0.393 |
| Dyslipidemia | 49 (44.5) | 13 (34.2) | 36 (50.0) | 0.167 |
| CKD | 3 (2.7) | 2 (5.3) | 1 (1.4) | 0.568 |
| Previous MI | 5 (4.5) | 3 (7.9) | 2 (2.8) | 0.457 |
| COPD | 6 (5.5) | 2 (5.3) | 4 (5.6) | 1.000 |
| Oxygen support |  |  |  | 0.299 |
| No/low flow | 61 (55.5) | 18 (47.4) | 43 (59.7) |  |
| High flow/NIV | 49 (44.5) | 20 (52.6) | 29 (40.3) |  |
| Steroid therapy | 110 (100.0) | 38 (100.0) | 72 (100.0) | 1.000 |
| Time from hospital discharge<br>to CPET (days) | 85.0 (71.5-102.5) | 81.0 (64.0-106.5) | 89.0 (78.3-98.5) | 0.463 |
| Length of hospital stay (days) | 18.0 (6.8-30.0) | 26.5 (15.0-33.8) | 15.5 (3.0-23.8) | 0.013 |
| **3-month clinical evaluation** |  |  |  |  |
| 6MWT (meters) | 540.0 (480.0-600.0) | 525.0 (480.0-567.5) | 540.0 (480.0-600.0) | 0.255 |
| Ejection fraction | 60.0 (60.0-60.0) | 60.0 (60.0-60.0) | 60.0 (60.0-60.0) | 0.485 |
| Dyspnea | 55 (50.0) | 19 (50.0) | 36 (50.0) | 1.000 |
| NYHA class III/IV | 18 (16.4) | 8 (21.1) | 10 (13.9) |  |
| Chest pain | 28 (25.5) | 9 (23.7) | 19 (26.4) | 0.937 |
| Angina pectoris | 2 (1.8) | 2 (5.3) | 0 (0.0) |  |
| Fatigue | 54 (49.1) | 21 (55.3) | 33 (45.8) | 0.459 |
| Palpitations | 25 (22.7) | 8 (21.1) | 17 (23.6) | 0.948 |
| Lipothymia/syncope | 2 (1.8) | 2 (5.3) | 0 (0.0) | 0.982 |

| Cardiopulmonary exercise testing |  |  |  |  |
| --- | --- | --- | --- | --- |
| Reason for CPET interruption |  |  |  | 0.654 |
| exhaustion/leg fatigue | 104 (94.5) | 34 (89.5) | 70 (97.2) |  |
| dyspnea | 4 (3.6) | 2 (7.9) | 2 (2.8) |  |
| new arrhythmia | 2 (1.8) | 2 (5.3) | 0 (0.0) |  |
| Peak VO2 (ml O2/min) | 1552.0 (1240.3-2068.0) | 1303.0 (1139.3-1542.3) | 1772.5 (1393.3-2259.8) | <0.001 |
| Percent predicted peak VO2 (%) | 90.9 (79.2-109.0) | 75.6 (64.8-79.2) | 99.2 (91.1-114.1) | <0.001 |
| Peak VO2/kg (mL O2/min /kg) | 20.6 (17.8-25.4) | 18.7 (15.2-20.5) | 22.9 (19.1-27.9) | <0.001 |
| Peak W (Watt) | 119.5 (92.0-167.5) | 101.5 (77.0-118.3) | 138.5 (104.0-187.8) | <0.001 |
| Percent predicted peak W (%) | 97.7 (70.7-113.6) | 61.2 (46.9-73.3) | 105.9 (95.5-120.2) | <0.001 |
| Peak HR (beat/min) | 149.5 (133.5-159.8) | 142.0 (115.5-158.8) | 150.0 (139.8-160.0) | 0.074 |
| Percent predicted HR (%) | 92.6 (84.7-98.1) | 88.7 (72.5-96.6) | 93.5 (88.8-98.8) | 0.012 |
| OUES | 93.5 (78.6-105.8) | 73.5 (65.7-80.1) | 99.4 (90.1-112.8) | <0.001 |
| RER | 1.1 (1.0-1.2) | 1.1 (1.0-1.2) | 1.1 (1.0-1.2) | 0.823 |
| VE (L/min) | 67.0 (54.9-88.3) | 62.8 (47.2-71.9) | 75.2 (60.5-96.6) | <0.001 |
| VT (mL) | 1754.0 (1457.3-2311.3) | 1596.5 (1391.8-1916.0) | 1940.0 (1535.8-2497.0) | 0.007 |
| BR (breath/min) | 39.0 (35.2-43.0) | 35.9 (32.1-41.9) | 39.4 (35.7-43.3) | 0.071 |
| BrR | 47.8 (28.1-67.4) | 60.9 (44.0-80.5) | 39.9 (21.8-52.5) | 0.002 |
| VE/VCO2 slope | 31.7 (27.8-34.7) | 33.2 (28.6-38.0) | 30.6 (27.7-33.3) | 0.041 |
| Maximal work rate (Watt) | 64.5 (44.0-95.5) | 54.0 (35.3-68.5) | 76.0 (50.8-103.5) | 0.001 |
| AT VO2 (mL O2/min) | 1054.5 (844.8-1382.5) | 883.5 (770.5-1111.5) | 1180.5 (962.0-1450.0) | <0.001 |
| AT HR (beat/min) | 112.0 (101.0-121.8) | 112.0 (93.3-121.0) | 111.5 (103.0-122.3) | 0.517 |
| AT VE/VCO2 | 33.6 (30.7-38.2) | 35.1 (31.0-41.1) | 33.3 (30.7-35.5) | 0.040 |
| VO2/HR (ml/beat) | 11.0 (9.1-14.2) | 10.0 (7.5-12.7) | 12.1 (9.8-15.5) | 0.001 |
| VO2/W slope | 9.1 (8.1-9.9) | 8.1 (7.0-8.9) | 9.5 (8.5-10.2) | <0.001 |
| Pulmonary function testing |  |  |  |  |
| FEV1 (L) | 3.0 (2.5-3.8) | 3.0 (2.6-3.5) | 3.0 (2.4-3.9) | 0.909 |
| Percent predicted FEV1 (%) | 104.0 (90.0-116.0) | 98.5 (87.0-109.8) | 108.0 (93.5-116.5) | 0.022 |
| FVC (L) | 3.6 (2.9-4.4) | 3.6 (3.2-4.1) | 3.6 (2.8-4.6) | 0.841 |
| Percent predicted FVC (%) | 100.0 (87.0-111.0) | 97.5 (85.3-104.5) | 102.0 (89.0-112.0) | 0.053 |
| Percentage of FEV1/FVC (%) | 106.0 (100.0-114.0) | 106.5 (98.8-110.0) | 106.0 (100.5-114.0) | 0.470 |
| MVV (L/min) | 120.0 (98.0-153.6) | 122.0 (104.8-140.7) | 118.4 (96.2-157.8) | 0.717 |
| PEF (L/s) | 7.3 (5.5-9.8) | 7.6 (5.3-9.2) | 7.0 (5.7-10.2) | 0.307 |
| Percent predicted PEF (%) | 82.0 (42.7-107.3) | 62.0 (40.0-103.8) | 87.0 (56.0-107.8) | 0.240 |
| FEF25-75% (L/s) | 3.1 (2.4-4.1) | 2.9 (2.5-3.9) | 3.1 (2.4-4.3) | 0.434 |
| MEF75% (L/s) | 6.3 (4.7-8.2) | 5.5 (4.6-7.5) | 6.5 (4.8-8.9) | 0.115 |
| MEF50% (L/s) | 4.0 (2.9-5.0) | 3.8 (2.8-4.6) | 4.0 (3.0-5.1) | 0.373 |
| MEF25% (L/s) | 1.4 (1.0-1.9) | 1.3 (1.0-1.8) | 1.43 (1.0-2.0) | 0.432 |
| Percent predicted DLCO (%) | 76.0 (65.5-95.3) | 70.0 (58.0-82.5) | 83.0 (69.0-103.0) | 0.063 |
| **Strength evaluation** |  |  |  |  |
| DLE maximal strength (kg) | 19.0 (12.0-29.3) | 17.0 (10.8-30.0) | 20.0 (12.4-27.0) | 0.201 |
| DLE maximal strength per BW | 0.3 (0.2-0.8) | 0.3 (0.2-0.4) | 0.2 (0.2-0.3) | 0.380 |

6MWT: 6 minutes walking test; AT: anaerobic threshold; BR: breathing rate; BrR: breathing reserve; CKD: chronic kidney disease; COPD: chronic obstructive pulmonary disease; CPET: cardiopulmonary exercise testing; DLCO: diffusing capacity of lungs for carbon monoxide; DLE: dominant leg extension; FEF25-75%: forced expiratory flow at 25-75% of FVC; FEV1: forced expiratory volume in one second; FVC: forced vital capacity. HR: heart rate; MEF: maximal expiratory flow; MI: myocardial infarction; MVV: maximal voluntary ventilation; OUES: oxygen uptake efficiency slope; Peak VO<sub>2</sub>: peak oxygen uptake; PEF: peak expiratory flow; RER: respiratory exchange ratio; VCO<sub>2</sub>: volume of exhaled carbon dioxide; VE: minute ventilation; VT: tidal volume; W: work level.

Forty-five (40.9%) patients were female, median age was 61.7 (53.5-69.2) years, median body mass index (BMI) at CPET evaluation was 26.8 (23.9-30.5) kg/m^2^, median weight at hospital admission was 82.0 (70.0-95.0) kg, whereas median weight at CPET evaluation was 77.0 (67.0 89.8) kg with median percent weight loss of 9.4 (6.0-12.9) %. Forty-five (40.9%) patients were active smokers.

### Cardiopulmonary exercise testing

The reason for maximal CPET interruption was exhaustion/leg fatigue for 109 (94.5%), dyspnea for 4 (3.6%), and new arrhythmia for 2 (1.8%). Median %pVO2 was 90.9 (79.2-109.0) %, median RER 1.1 (1.0-1.2). Thirty-eight (34.5%) patients had %pVO2 below, whereas 72 (65.5%) above the 85% predicted value.

Of the 38 patients with reduced %pVO2, 8 (21.1%) had mainly respiratory limitation of exercise (RLE, *see online-only material for definitions*), 9 (23.7%) mainly cardiac limitation of exercise (CLE), 3 (7.9%) RLE and CLE, and 18 (47.4%) had non-cardiopulmonary limitation of exercise.

### Pulmonary function test

At PFT, median forced expiratory volume in one second (FEV1), forced vital capacity (FVC), and diffusing capacity of lungs for carbon monoxide (DLCO) were within normal limits; however, 2 (1.8%) patients had mild and 2 (1.8%) moderate impairment of FEV1, 1 moderate impairment of FEV1, 5 (4.5%) mild and 2 (1.8%) impairment of FVC, and 9 (8.2%) mild, 6 (5.5%) moderate and 2 (1.8%) severe DLCO impairment.

### 3-month clinical evaluation

At 3-month clinical evaluation, 81 (73.6%) patients experienced at least one disabling symptom, 30/38 (78.9%) among those with %pVO2 below and 51/72 (70.8%) among those above 85% (p>0.05). Of note, 55/110 (50.0%) patients complained of dyspnea, of whom 18/110 (16.4%) were in NYHA class III/IV, 28/110 (25.5%) had chest pain, 54/110 (49.1%) had fatigue, and 25/110 (22.7%) complained of palpitations. Each symptom frequency did not differ between patients with %pVO2 below and above 85% predicted value (all p>0.05).

### Predictors of percent predicted peak oxygen uptake (VO2)

At multivariate linear regression analysis adjusted for time from hospital discharge to CPET, age (β=0.46, p=0.020), percent weight loss (β=-0.77, p=0.029), active smoke status (β=-7.07, p=0.019), length of hospital stay (β=-0.20, p=0.042), and DLE maximal strength (β=1.65, p=0.039) (**Figure 1**) were independently associated with %pVO2 (**Table 2**).

**Table 2.** **Univariate and multivariate linear regression for percent predicted oxygen uptake (VO2) adjusted for time from hospital discharge to cardiopulmonary exercise testing.**

| Variable | Univariate |  |  | Multivariate |  |  |
| --- | --- | --- | --- | --- | --- | --- |
| | $\beta$ coefficient | 95%confidence interval | p value | $\beta$ coefficient | 95%confidence interval | p value |
| **Baseline and clinical characteristics** |  |  |  |  |  |  |
| **Age (years)** | **0.58** | **0.16 to 1.00** | **0.007** | **0.46** | **0.07 to 0.85** | **0.020** |
| Sex (female) | -4.28 | -14.94 to 6.39 | 0.426 |  |  |  |
| Height (centimeters) | 0.08 | -0.46 to 0.63 | 0.758 |  |  |  |
| Absolute weight loss (kg) | -1.33 | -2.37 to -0.29 | 0.013 |  |  |  |
| Weight at CPET evaluation (kg) | 0.37 | 0.03 to 0.71 | 0.035 |  |  |  |
| Weight at hospital admission (kg) | 0.16 | -0.15 to 0.47 | 0.297 |  |  |  |
| **Percent weight loss (%)** | **-1.33** | **-2.29 to -0.37** | **0.007** | **-0.77** | **-1.58 to -0.44** | **0.029** |
| Body mass index (kg/m<sup>2</sup>) | 0.81 | -0.07 to 1.70 | 0.071 | 0.66 | -0.27 to 1.59 | 0.158 |
| Hypertension | 4.43 | -6.53 to 15.38 | 0.423 |  |  |  |
| Diabetes | -27.74 | -58.66 to 3.19 | 0.178 |  |  |  |
| **Active smoke** | **-9.17** | **-14.04 to -3.69** | **0.037** | **-7.07** | **-16.75 to -2.60** | **0.019** |
| Dyslipidemia | -1.33 | -12.14 to 9.49 | 0.807 |  |  |  |
| CKD | -47.08 | -90.00 to 4.16 | 0.102 |  |  |  |
| Previous MI | -4.50 | -24.97 to 15.97 | 0.662 |  |  |  |
| COPD | -6.03 | -28.72 to 16.67 | 0.598 |  |  |  |
| Oxygen support (high flow/NIV) | -9.71 | -20.22 to 0.79 | 0.169 |  |  |  |
| **Length of hospital stay (days)** | **-0.34** | **-0.64 to -0.03** | **0.031** | **-0.20** | **-0.73 to -0.03** | **0.042** |
| **3-month clinical evaluation** |  |  |  |  |  |  |
| 6MWT (meters) | 0.02 | -0.05 to 0.06 | 0.893 |  |  |  |
| Ejection fraction | 1.03 | 0.05 to 2.01 | 0.039 | 0.22 | -0.69 to 1.14 | 0.630 |
| Dyspnea (NYHA class III/IV) | -4.51 | -15.19 to 6.18 | 0.403 |  |  |  |
| Chest pain | -2.84 | -43.21 to 33.21 | 0.642 |  |  |  |
| Weakness | -11.48 | -21.83 to -1.14 | 0.030 |  |  |  |
| Palpitations | -7.77 | -19.75 to 4.20 | 0.199 |  |  |  |
| Lipothymia/syncope | -3.11 | -47.56 to 41.33 | 0.889 |  |  |  |
| **Pulmonary function test** |  |  |  |  |  |  |
| Percent predicted FEV1 (%) | 0.33 | 0.09 to 0.58 | 0.007 | 0.21 | -0.01 to 0.44 | 0.066 |
| Percent predicted FVC (%) | 0.29 | 0.03 to 0.56 | 0.029 |  |  |  |
| Percentage of FEV1/FVC (%) | 0.46 | -0.03 to 0.94 | 0.066 |  |  |  |
| Percent predicted PEF (%) | 0.01 | -0.13 to 0.15 | 0.839 |  |  |  |
| Percent predicted DLCO (%) | 0.22 | -0.08 to 0.51 | 0.144 |  |  |  |
| **Strength evaluation** |  |  |  |  |  |  |
| **DLE maximal strength (kg)** | **1.99** | **1.10 to 2.87** | **0.030** | **1.65** | **1.16 to 1.87** | **0.039** |

6MWT: 6 minutes walking test; BW: body weight; CKD: chronic kidney disease; COPD: chronic obstructive pulmonary disease; DLCO: diffusing capacity of lungs for carbon monoxide; DLE: dominant leg extension; FEF25-75%: forced expiratory flow at 25-75% of FVC; FEV1: forced expiratory volume in one second; FVC: forced vital capacity. MI: myocardial infarction; PEF: peak expiratory flow.

**Figure 1.**
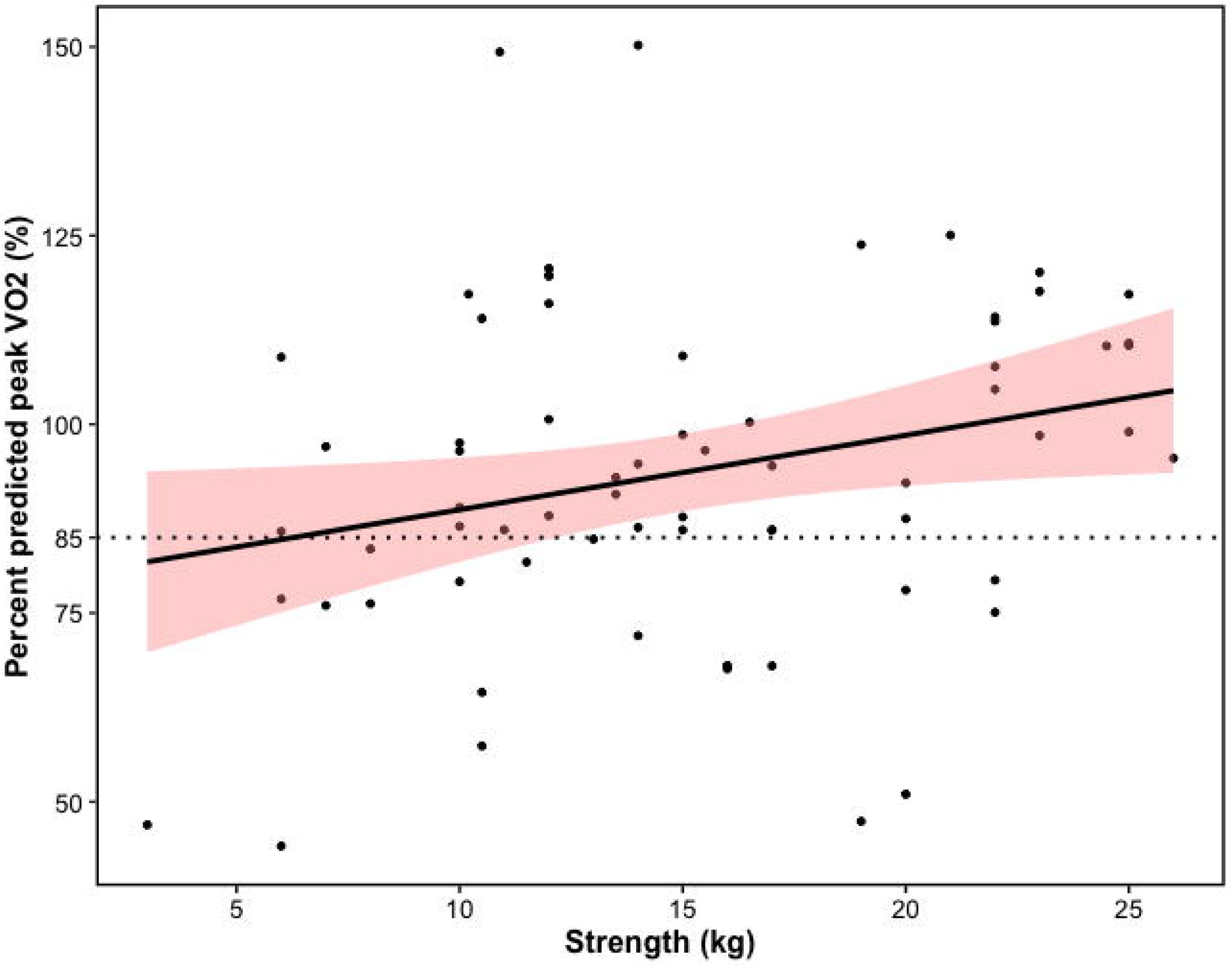
Percent predicted peak oxygen uptake (VO2) per dominant leg extension strength.

## DISCUSSION

Our study has the following main findings: 1) 1/3^rd^ of non-severe COVID-19 survivors had a significant alteration both in exercise capacity and %pVO2 at 3 months after hospital discharge; 2) in about half of patients with abnormal %pVO2, this was due to abnormal peripheral oxygen extraction, most likely to some degree of muscle impairment, as DLE maximal strength was independently associated with peak oxygen consumption; 3) more than 2/3^rd^ (74.3%) of patients experimented at least one disabling symptom at 3 months after hospital discharge, although there was no relationship between symptoms and worst %pVO2.

To our knowledge, for the first time we assessed clinical status and exercise capacity of COVID-19 patients performing complete CPET evaluation after hospital discharge.

Regarding the first point, our results are reminiscent of Ong et al., who found a 41% prevalence in reduced %pVO2 among 44 SARS 3-month survivors[8], albeit they also included 10/44 (22.7%) patients that had required invasive ventilation. The abnormal %pVO2 was accompanied by an early anaerobic threshold, translating into a significant impairment in daily activities.

On note, abnormal physical function and performance in COVID-19 survivors have been preliminarily described by Belli et al.[9] using 1-min sit-to-stand test and Short Physical performance Battery, without the more objective CPET evaluation.

Due to the ongoing and accelerating COVID-19 worldwide pandemic, these observations raise important concerns for health systems, as we proved that a substantial number of non-severe COVID-19 patients still had objective exercise impairment several months after hospital discharge. As for the second point, it is noteworthy that a cardiopulmonary cause determining the exercise capacity and %pVO2 reduction could only be found in about half of patients.

Interestingly, DLE maximal strength was independently associated with peak oxygen consumption suggesting that muscle impairment should be responsible for the residual cases, probably due to bed rest and subsequently muscular deconditioning, but also with a potential role for corticosteroid myopathy[10, 11]. Our numbers are again similar to Ong et al. (about 40% had non-cardiopulmonary impairment in their post-SARS cohort), although steroids during acute illness was less frequently used in their sample (15% vs. 100%). The important role of muscular factors is compounded by the higher weight loss in patients with abnormal %pVO2, probably representing a reduction in fat and lean mass occurring during the acute phase.

We also highlight the role of hospital stay, that appeared longer among patients with abnormal %pVO2, probably for both the forced confinement during the acute phase and the need for aggressive medical care in COVID-19. Regarding this, our data can again only be compared with Belli et al.[9], which found no association between physical performance and length of hospital stay, but without measuring objective CPET-derived parameters.

We believe that the most important of our findings is the relationship between %pVO2 and maximal strength of the lower limb muscles, maintained even after accounting for cardiopulmonary variables, for length of hospitalization, and for percent weight loss. In our opinion this reduces the likelihood that bed rest, exercise deprivation and loss of muscle mass alone could cause this degree of impairment, raising the possibility of a direct effect of SARS-CoV2 at the muscle level. We only mention the possibility of mitochondrial dysfunction[12], as several ongoing research projects are exploring its role on the pathogenesis of COVID-19 acute phase[13-15].

As for the third point, we demonstrate that almost 3/4^ths^ of patients experienced at least one disabling symptom 3 months after hospital discharge, without any relationship with exercise capacity. Several studies have already investigated the residual symptoms burden of patients recovering from COVID-19, observing different rates between out-patients (about 35% in Tenforde et al.)[16] and patients who had needed hospitalization (87% in the study by Carfì et al.)[6]. We report relatively high rates of disabling symptoms (50% dyspnea, 49.1% fatigue). Halpin et al. describe a cohort of 68 non-ventilated patients, of whom 41/68 (60%) complained of fatigue and 21/68 (43.6%) of dyspnea. The need for use of oxygen supplementation (80% vs. 70% in Halpin et al.) was also similar [17].

In conclusion, it has been known for many years that most critically ill patients face long-lasting functional impairment after discharge[10]; what is mostly worrying about our data is that we found severe mid-term consequences of COVID-19 in a non-ICU population. This observation supports the need for targeted management of these patients also during the acute phase (e.g. applying appropriate nutrition and early mobilization plans). Moreover, as there was no relationship with %pVO2, symptoms alone should not guide the post-acute management of COVID-19 patients: more objective techniques, such as CPET, should probably be used to rapidly intercept and assess the exercise impairment and, perhaps, to decide whether to start a physical rehabilitation program.

### Limitations

Our study has important limitations. Firstly, all patients came from an area of the city of Genoa. Secondly, the functional capacity evaluation was conducted three months after hospital discharge, with the patients unsupervised in the meantime and no data available about the baseline condition prior to COVID-19. Moreover, no direct structural evaluation at the muscle level was performed.

## Data Availability

All data are available upon request.

## ACNOWELDGEMENTS

None.

## Notes

### Competing Interest Statement

The authors have declared no competing interest.

### Author Declarations

All patients signed an informed consent. Study protocol and informed consent conform to the Declaration of Helsinki and were approved by the Ethics Committee of the Liguria Region (n 430/2020CER).

