## Supplemental material and methods for "Assessment of functional capacity with cardiopulmonary exercise testing in non-severe COVID-19 patients at three months follow-up"

**Cardiopulmonary exercise testing**

Cardiopulmonary exercise testing (CPET) was performed according to European Respiratory Society (ERS) standard criteria[1] and using the COSMED system for cardiopulmonary exercise testing (Quark CPET, COSMED, Rome, Italy) with an electronically braked cycle ergometer (Excalibur Sport, Lode, Groningen, Germany) under the supervision of a physician. The height of the bike seat was adjusted to keep the subject legs at near the full extension during each pedal revolution.

One of four possible ramp protocol was selected (10, 15, 20, 25 watt/min) based on sex and weight in order to reach the maximal possible oxygen consumption in 10 to 12 minutes. Encouragement to maintain a pedaling rate between 60 and 70 revolution/minute continued throughout the duration of each study.

The ramp protocol started after 5 minutes of rest on the bike and 2 minutes of unloaded pedaling. Standard 12-lead electrocardiograms and blood pressure measurements were obtained at rest, every 3 minutes during exercise, at peak exercise and for 5 minutes into the recovery phase.

The gas analyzer was calibrated for accuracy and linearity prior to each test in accordance with the manufacturer’s instructions. Metabolic measure were obtained by the breath-by-breath system throughout the study.

End of test criteria where met when patients were unable to maintain a pedaling cadence above 60 revolutions/minute. The test was considered maximal if the respiratory exchange ratio (RER) was above 1.1. and/or if peak heart rate was above 85% of predicted maximal heart rate.

Oxygen uptake (VO2; measured in mL/minute; standard temperature and pressure, dry), carbon dioxide production (VCO2; measured in mL/minute), gas exchange ratio, maximum exercise ventilation (VE; measured in L/minute; at body temperature, ambient pressure and saturated with water vapor), breathing rate (BR), tidal volume (VT) and the ventilatory equivalent for carbon dioxide (VE/VCO2) were determined and averaged every 30 seconds.

Breathing reserve (BrR) was defined as either the difference between the maximal voluntary ventilation (MVV) and the VE in absolute terms, or this difference as a fraction of the MVV.

The peak oxygen uptake (peak VO2) was defined as the highest 20-seconds averaged VO2 during the last minute of the symptom limited exercise test.

Maximal work rate was defined as the highest work level that was reached. The anaerobic threshold (AT) was identified for each subject using both the V-slope and ventilator equivalent methods. Aerobic capacity reduction was defined as when peak VO2 was 85% below the predicted value. The Hansen and Wasserman equation was used to determine the normal predicted maximal O2 value[2].

The interpretation of CPET was done independently by two experienced cardiopulmonary exercise testing researchers; when the interpretation was discordant, conclusive third evaluation was done by a senior CPET researcher.

**Mechanisms of peak Vo2 reduction**

Exercise capacity and aerobic capacity limitations are always multifactorial: then single measurements are not sufficient to determine a main reason to justify a reduced exercise capacity. Various algorithms have been proposed to guide the clinical interpretation of CPET but with insufficient clinical validation. For these reasons we decided to adopt an integrated interpretation approach to identify, if possible, a “main” reason causing the exercise limitation in every single patient even according ERS recommendation for CPET[1].

Cardiovascular limitation of exercise (CLE) was defined when one or more of the following criteria were present: a history of chronic heart conditions, evidence of cardiac systolic or diastolic disfunctions, reduced exercise capacity with early appearance of lactate threshold, reduced oxygen pulse with relatively flat progression during exercise, maximal heart rate close to the maximum predicted.

Respiratory limitation of exercise (RLE) was defined when one or more of the following criteria were present: a history of chronic lung disease, a significant reduction of pulmonary function at rest as FEV1 <70% of predicted and/or DLCO <70%. A BrR <15% at peak of exercise was also considered as a cut-off value for respiratory limitation of exercise.

In some cases, the coexistence of cardiac and respiratory factors was indicated as a limiting factor.

In absence of cardiac or respiratory limitation and/or in presence of leg fatigue as a limiting factor of exercise also in presence of normal breathing and heart rate reserve, the VO2 reduction was interpreted as a consequence of abnormal systemic oxygen extraction. This could be the consequence of different mechanisms but the muscular hypothesis level seem to be the more plausible because of the CPET parameters pattern (reduced peak VO2, reduced AT VO2, reduced peak HR, VE/MVV<85% and normal SpO2)[1], because of the frequent muscular pain complain in the post COVID-19 patients medical history, because of the lack of muscular strength and resistance and because of the dramatic weight and lean mass loss during the acute phase of the disease.

The indication of the main reason for limiting the exercise capacity was done independently by two experienced cardiopulmonary exercise testing researchers; when the interpretation was discordant, a third evaluation by a senior CPET researcher was performed.

Oxygen saturation via pulse oximetry and HR by electrocardiography were recorded continuously throughout exercise and during recovery. At the end of exercise, the main reasons for termination were obtained from the subjects.

**Pulmonary function testing**

Pulmonary function test (PFT) included spirometry and measurements of diffusing capacity of lungs for carbon monoxide (DLCO). Spirometry was performed in accordance with recommended standards. All lung function tests were performed with the subject seated and on the same day, but before the exercise test. Forced vital capacity (FVC) and forced expiratory volume in one second (FEV1) were measured with a clinical spirometer (Bodystik by Geratherm, Germany). DLCO was determined by the single-breath carbon monoxide technique using an infrared analyser (Diffustik by Geratherm, Germany), which utilises methane as an inert tracer gas.

DLCO was adjusted for the hemoglobin concentration evaluated on the day of the PFT. The PFT and DLCO measurements were expressed as percentages of predicted normal values using reference values taken from prediction equations previous described[3, 4].

**Dominant length extension evaluation**

Muscle strength (kg) was tested with air resistance equipment (HUR Oy, Kokkola, Finland), which replaces the weight plates traditionally used in weight stack machines with a pneumatic system of resistance. Maximal dynamic strength of leg extension from the dominant side was assessed to find the maximum load with which a subject was able to perform 3-5 repetitions. From these loads the expected 1-repetition maximum (1-RM) was estimated by using Brzycki's formula integrated to HUR SmartTouch software[5].
